# Study Protocol for Developing a Public Health Registry: The Flint Registry Experience

**DOI:** 10.1101/2025.08.25.25332399

**Authors:** Nicole Jones, Marty Crawford, Jenny LaChance, Kenyetta Dotson, Mona Hanna

**Affiliations:** Michigan State University–Hurley Children’s Hospital Pediatric Public Health Initiative, Charles Stewart Mott Department of Public Health, Michigan State University College of Human Medicine, Flint, Michigan 48502, USA; Department of Pediatrics and Human Development, Michigan State University College of Human Medicine, East Lansing, Michigan 48502, USA

**Keywords:** Flint water crisis, Lead, Public health registry, Community engagement, Public health surveillance, Secondary prevention, Public health authority

## Abstract

**Background:** From April 25, 2014, to October 15, 2015, residents of Flint, Michigan, were unknowingly exposed to contaminated water after a drinking water source switch. This public health disaster, the Flint water crisis (FWC), resulted in population-wide lead-in-water exposure for 18 months. A potent neurotoxin, lead is associated with multiple long-term adverse health conditions. In response to the FWC, the Flint Registry (FR) was formed to mitigate the impact of the crisis by proactively identifying the health and development concerns of participants and providing secondary prevention resources to address their needs.

**Methods:** Designed as a public health intervention with robust community engagement in every aspect, the FR enrollment is non-end-dated with ongoing evaluation and screening. Eligibility requirements include individuals who lived, worked, or went to school at an address serviced by the City of Flint water system anytime from 4/25/2014 to 10/15/2015, including those exposed prenatally. Recruitment is list-based, community-outreach-based, and marketing-based. Participants enroll primarily online or by phone and complete a survey to evaluate health impacts of the FWC and screen for service needs. Data collected at enrollment includes demographics, physical/mental health, child development, prior utilization of services, and environmental/lead-exposure risks. Based on responses, enrollees are referred within a community referral network to services in the categories of lead elimination, health, nutrition, and child development via an automated process. Frequencies of reported diagnoses, health symptoms, food access/insecurity problems, and educational support needs are compared to city, county, state, and national measures to identify ongoing disparities. Referrals identify the unmet needs for secondary prevention services.

**Discussion:** The FR protocol is unique because it not only conducts longitudinal surveillance, but it also improves public health through a community-wide referral process, allowing many people to be connected to health-promoting resources. This protocol is applicable to other public health crises due to its broad, city-wide surveillance and mitigation efforts, reduction of barriers to maximize participation, and incorporation of community voice. The FR protocol was designed in concert with the community, and community engagement remains a priority. Future work includes a diversified community engagement approach, ongoing enrollment, referrals, surveillance, and mitigation efforts.

## BACKGROUND

On April 25, 2014, the source of the city of Flint’s water was switched from Lake Huron via the Detroit Water and Sewerage Department to the Flint River via the Flint Water Treatment Plant. The cost-cutting decision was made by anti-democratic, state-appointed financial emergency managers [1]. An environmental and public health injustice, Flint residents unknowingly consumed lead-contaminated water for 18 months (April 25, 2014, to October 15, 2015). The impacts of the Flint water crisis (FWC) are many and include a population-wide increase in water lead levels and childhood blood lead levels, along with a decrease in public trust and increase in mental health concerns [2–5].

A potent neurotoxin with no safe level, exposure to lead is associated with neurologic, renal, cardiovascular, hematologic, reproductive, and developmental problems [6]. Further, the toxic stress model suggests that trauma caused by chronic social conditions or accumulating adverse events in the absence of social support systems can impair the development of the nervous system and the ability to self-regulate emotions, overall cognitive performance, and both physical and mental health [7,8]. Toxic stress affects the body and brain throughout the life-course but is especially detrimental in early childhood because of the cumulative life-long impacts. Under this model, the injustice of the FWC is an added stressor atop multiple pre-existing health-harming systemic inequities (ie: poverty, racism, unemployment, etc.) within the Flint community.

In response to the FWC, public health infrastructure was expanded to mitigate the potential adverse impact on health. The Flint Registry (FR) is an ongoing project that was modeled after other exposure registries such as the World Trade Center Health Registry [9] but is unique in that it conducts surveillances and includes an expansive service referral component to improve public health.

### Formative Work

Shortly after the uncovering of the city-wide lead exposure (September 2015), there was recognition of the need for an exposure registry to identify and longitudinally support those impacted by the crisis. The creation of a registry was included in recommendations submitted to the Emergency Operations Center during the federal declaration of emergency (January 2016) (see Figure 1: Timeline for launch of Flint Registry). Additionally, a “toxic registry” was recommended in the Flint Water Advisory Task Force Final Report in March 2016 [1]. Local partners from Flint-based Michigan State University (MSU) and Greater Flint Health Coalition (GFHC) began to plan for a registry and worked with the local health system to add a “FWC exposure” to the diagnosis list of applicable patient electronic medical records at Hurley Medical Center and with the Michigan Department of Health and Human Services (MDHHS) to add a similar indicator to the Michigan Care Improvement Registry for applicable children. In January 2017, MSU received a FR planning grant from MDHHS [10] to establish the leadership team, investigate informatics solutions, identify and strengthen community partnerships, and create an overall workgroup structure to design and implement the FR. The FR leadership team consisted of the registry director and principal investigator at MSU, the CEO of the GFHC, and the leadership of the City of Flint. Workgroups (Figure 2) with multi-disciplinary expertise were created to provide expertise in topics including: community lead elimination strategies, public health law, communications, service referrals, survey methodology, informatics, community outreach and engagement, and evaluation. Community representation was prioritized in formation of the workgroups. After congressional approval, the Centers for Disease Control and Prevention (CDC) released the Notice of Funding Opportunity (May 2017) and awarded the grant (August 2017) to MSU to continue planning and implementing a Flint Lead Exposure Registry. To facilitate the work of the FR, the CDC granted public health authority to the FR at MSU [11]. This allowed MSU to receive recruitment and health outcomes data from other public health agencies for the purposes of responding to the public health emergency.

**Figure 1.**
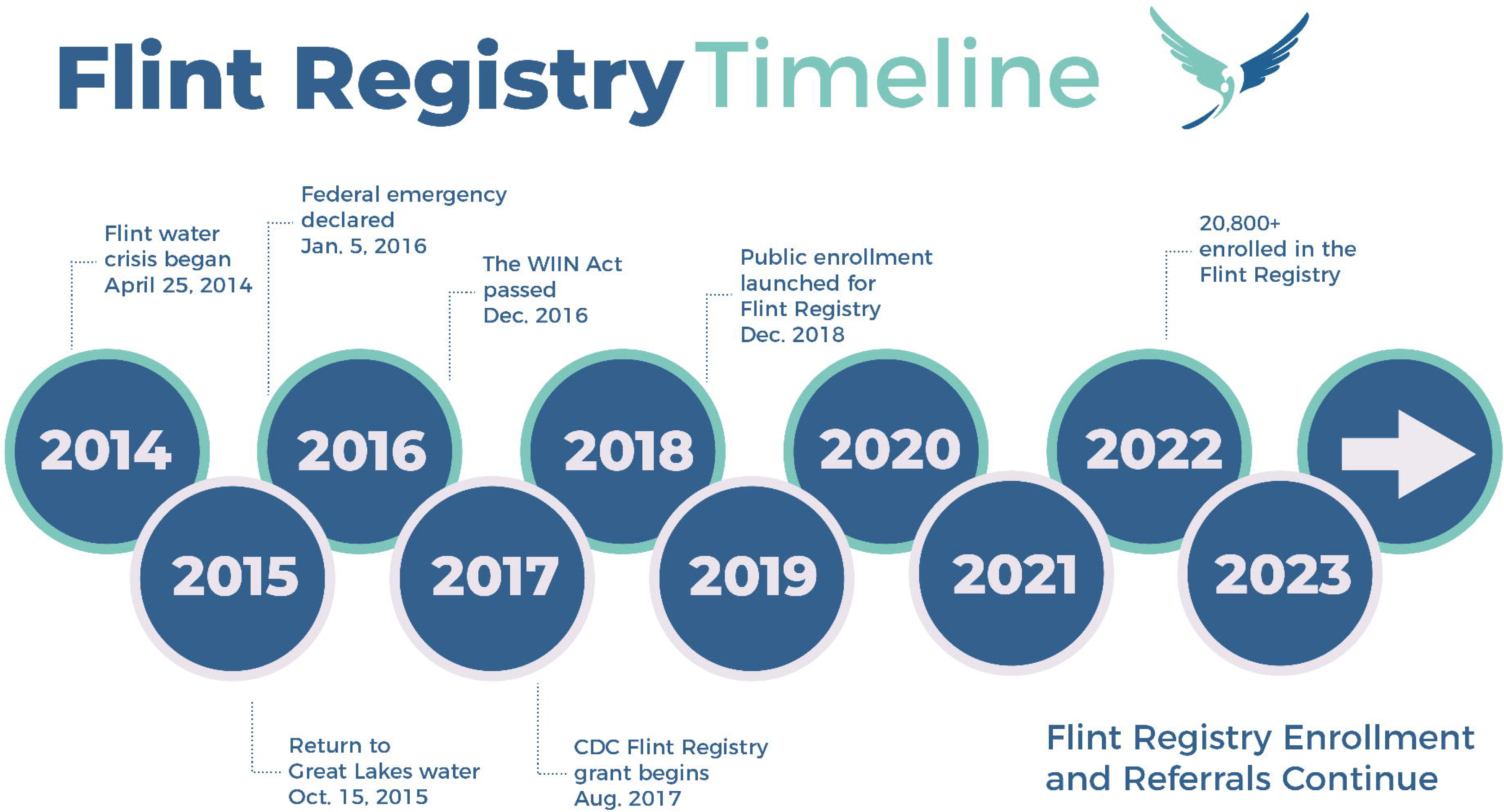
Timeline for Launch of Flint Registry.

**Figure 2.**
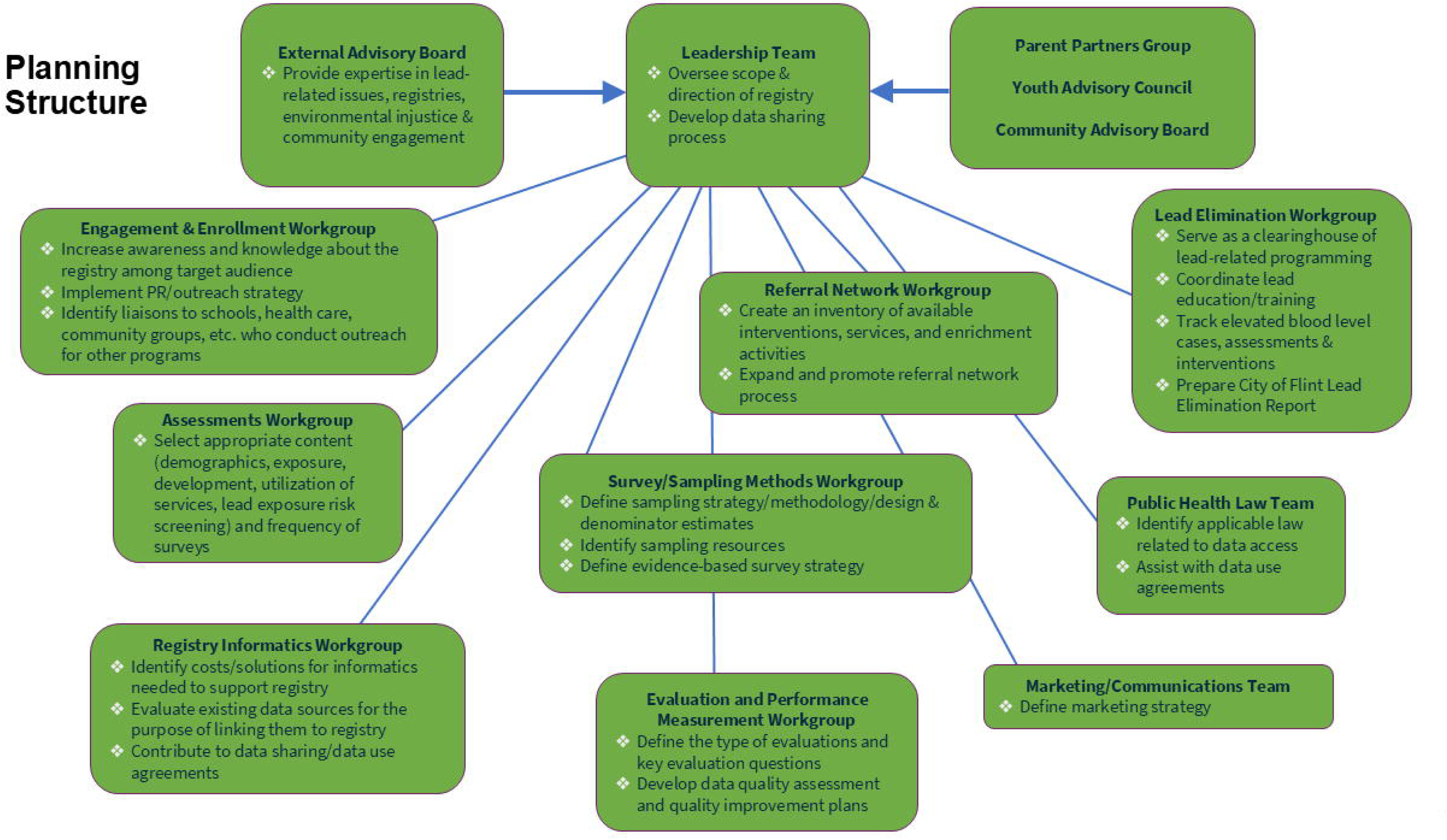
Flint Registry Planning Workgroups.

### Stakeholder Engagement

A foundational philosophy and practice of the FR has been the deliberate inclusion and engagement of community members as co-creators of knowledge, from inception and launch to its current format. Community engagement strategies included partnering with community-based organizations, hosting community events, collecting pre-enrollment feedback, conducting focus groups, community conversations, developing a community ambassador program, hiring and training community members, creating and funding a position of Director of Community Engagement and Implementation, and convening advisory boards. The Flint Registry Community Advisory Board (CAB) has a mission to increase FR community awareness, to facilitate input, education and support, and to coordinate activities to benefit registrants. The CAB includes a diverse mix of residents, organizational stakeholders, and young adults from key sectors of the Flint community: faith, workforce development, seniors, education, academia, organized labor, non-profit, government, philanthropy, media, health care, parents, youth, and law enforcement. CAB members have been and remain key informants of community voice regarding marketing and outreach, data collection methods, and outcomes reporting. A FR external advisory board with national experts in environmental injustice, lead and health, public health registries, and pediatric health was also convened to assist with planning. The MSU and Hurley Children’s Hospital Pediatric Public Health Initiative (PPHI) has two additional community advisory groups that also informed FR implementation: 1) Parent Partners, which includes parent representation from all nine wards of the city of Flint, and 2) Flint Youth Justice League, which includes Flint children, ranging in age and demographics. Lastly, as an effort to engage with already-enrolled participants, especially to inform longitudinal surveillance and cohort maintenance, the FR invited participants to join a FR Member Advisory Council. We will continue to engage with all advisory groups to identify additional outreach and recruitment strategies and to inform content and dissemination strategies of FR outcomes/results. Full details of the community engagement strategy were previously described [12].

### Project Aims and Hypothesis

The specific aims of the FR project are 1) to identify health and development concerns and capture the real-life, lived experiences of families impacted by the FWC and 2) to identify and address service needs and provide secondary prevention resources for participants through connection to an extensive referral process. The FR will also create the infrastructure for long-term surveillance and support, as well as multi-sector lead elimination work via the FR workgroup Flint Lead Free. It is anticipated that enrollees in the FR will have a higher prevalence of lead-and crisis-associated health and child development conditions as compared to other county, state, and national data. Service needs are supported in the areas of lead abatement, health services, nutrition, and child development programming. It is hypothesized that enrollment in the FR is expected to increase utilization of preventative services and, thus, improve health and development.

## METHODS/DESIGN

### Study Design

The FR protocol is designed as a public health intervention for persons exposed to the FWC to mitigate the long-term impact of lead exposure by increasing individuals’ use of health-promoting secondary prevention programs and services. Individuals who enroll in the registry complete a survey designed to assess their health and development and, based on their responses, are referred to high-priority, health-promoting services. Figure 3 displays the steps from recruitment to referral to data analysis. Because the manifestations of lead and trauma exposure may develop over years and decades [13,14], the FR supports ongoing enrollment, referrals, and evaluation. The FR goal is to enroll a minimum of 20% of the eligible population and at least 25% of groups with a higher risk of exposure or adverse outcomes. The MSU Institutional Review Board reviewed the FR protocol and determined it did not meet the definition of “research” as defined by the U.S. Department of Health and Human Services regulations for the protection of human research subjects. In addition, the FR received an endorsement from the local volunteer-led Community Based Organization Partners Community Ethics Review Board (CERB) after a complete review which determined the Flint Registry project to be “ethically sound and consistent with the vision and mission of the CERB.” This project was conducted in accordance with the ethical principles and guidelines for the protection of human subjects of research as described in Belmont Report prepared by United States National Commission for the Protection of Human Subjects of Biomedical and Behavioral Research in 1979.

**Figure 3.**
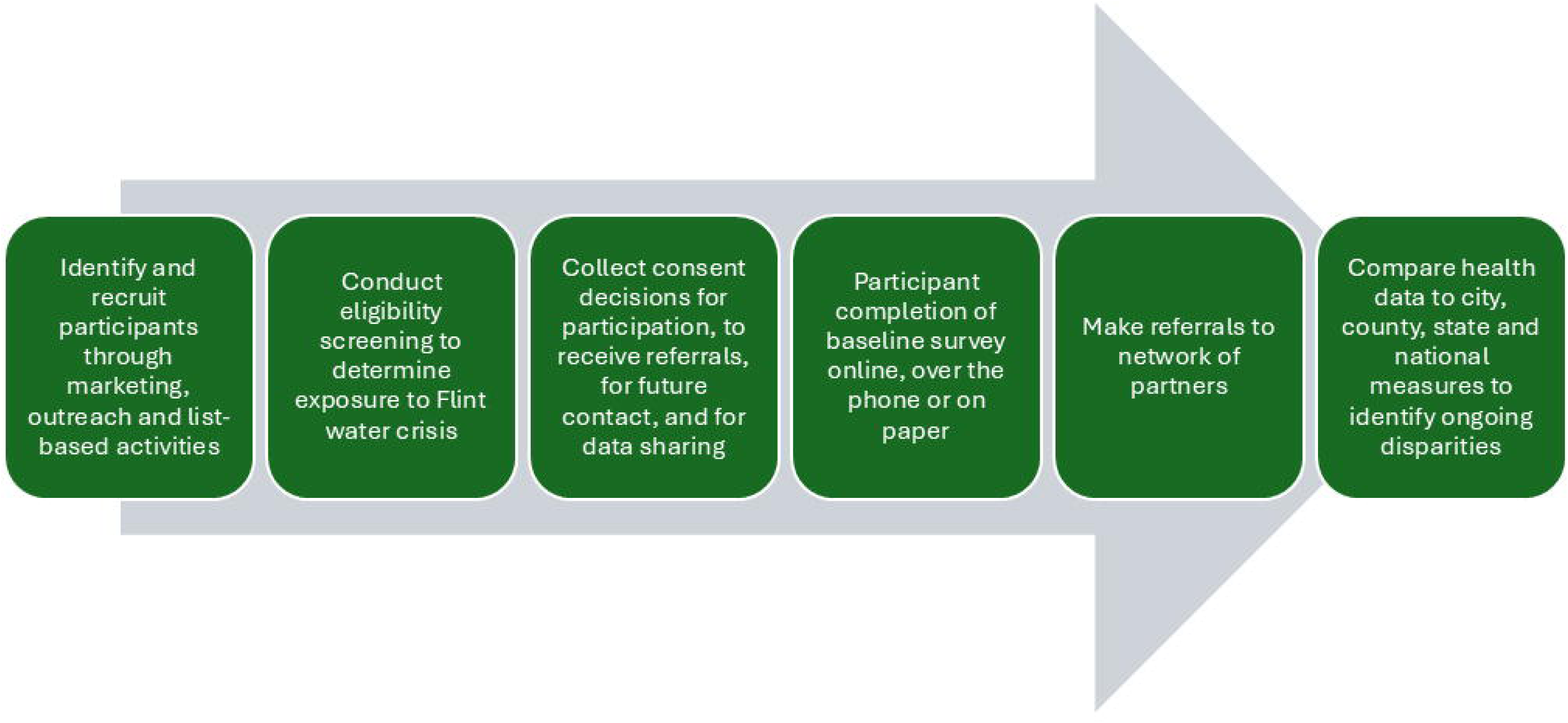
Fint Registry Protocol.

### Project Setting

The project setting is the city of Flint, Michigan, and is focused on exposure to the Flint water system during the period from April 25, 2014, to October 15, 2015. During this time, Flint’s water was switched from the Great Lakes to the Flint River and treatment of the water lacked adequate corrosion control. Community-informed planning began in 2016, and recruitment and enrollment began in 2018 and is ongoing with no planned end-date. Individual resident characteristics and overall household characteristics were used to develop recruitment strategies and to understand environmental health factors. Census data at the time of the FWC (2014-2015) were used to estimate the number of exposed individuals, to calculate the number of individuals with higher risk for health impacts of the FWC, to understand the prevalence of factors related to ongoing exposure to lead, and to assess potential eligibility for services.

Census data at the time of project launch (2018) describe the sociodemographics of the community. With respect to age, one-quarter of Flint residents are under 18 [15]. Residents have a lower level of education and income and higher rates of poverty as compared to Michigan residents. Among Flint residents over age 25, 85% have at least a high school diploma and 12% have a Bachelor’s degree or higher, as compared to 91% of State of Michigan residents with at least a high school diploma and 29% having a Bachelor’s degree or higher [16]. The median household income is $27,717 with 40% of persons and 58% of children living in poverty, compared to a state-wide median income of $54,938 and 15% of persons and 21% of children living in poverty [17]. The most common form of health insurance is public insurance with 66% of residents on public insurance [17]. English is the primary language and is the only language spoken in 96.4% of households [16]. However, Spanish is the primary language in 2.1% of households [16], and although not measured in the census, Flint also has a significant number of residents who communicate using American Sign Language and is the home of the Michigan School for the Deaf. City housing stock is older (92% built prior to 1980), slightly majority owner-occupied (55%), and representative of a shrinking city (25% vacant) [18].

### Participants

#### Eligibility criteria

FR eligibility criteria are based on the CDC Notice of Funding Opportunity [11] and include individuals who lived, worked, or went to school at an address serviced by the City of Flint water system anytime from 4/25/2014 to 10/15/2015, including those exposed in utero. Based on these eligibility criteria an estimated denominator was calculated, high-risk groups were identified, and data sources for recruitment were identified.

#### Denominator estimate

The FR sampling workgroup, with experts in statistics, geospatial analysis, epidemiology, informatics, and survey methods, estimated the number of exposed individuals (denominator) and identified methods to recruit from the sampling frame. Denominator estimates were calculated for the year 2014 and focused on four groups: 1) city of Flint residents, 2) those residing outside of Flint who worked within the city of Flint, 3) residents living outside of the city boundary whose homes were serviced by the Flint municipal water system, and 4) children attending school within the city of Flint who lived outside the city of Flint (Table 1). Data sources were not readily available to estimate the number of individuals who lived outside the city of Flint but attended daycare within the city of Flint.

**Table 1.** Estimated number of individuals eligible for the Flint Registry by category (2014)

| Category | Eligible Population Estimate |
| --- | --- |
| City of Flint Residents | 100,569 |
| Non-City of Flint Residents Serviced by the Flint Water System | 430 |
| Flint Employees Residing Outside the City | 32,575 |
| Flint Students Residing Outside the City | 1,214 |
| Total Denominator Estimate | 134,788 |

The number of residents living in the city of Flint during the eligibility period was calculated using the American Community Survey (ACS) 5-year estimates obtained from American FactFinder [19]. The number of people living outside and working inside the city of Flint was estimated using “OnTheMap”, a tool hosted by the U.S. Census Bureau that uses Longitudinal Employer-Household Dynamics (LEHD) Origin Destination Employment Statistics (LODES) [20]. The estimated number of non-city of Flint residents who used Flint city water was based on a list of addresses released by the City of Flint [21]. Each address was geocoded, and a 2017 parcel layer from the Genesee County GIS Department was used to omit commercial and vacant addresses. The remaining addresses were summed within each block group and multiplied by the estimated number of people per household (determined using ACS data). The population of students living outside of Flint who attended Flint schools or who attended the one school located outside of the city boundary and serviced by the City of Flint water system was estimated by Genesee Intermediate School District (GISD) student home address records. Each type of denominator estimate was calculated in such a way as to eliminate population overlap. However, a small number of people who lived outside the city and who also worked or attended school in Flint may have been represented in more than one denominator. This potential amount of overlap, no more than 0.34% of the total denominator estimate, was considered negligible. The total number of individuals in the 4 denominator categories was 134,788. Of the four denominator categories, Flint residents (approximately 100,000) were given highest priority for recruitment, and a recruitment goal of 20,000 residents (20%) was selected based on response rates reported by other voluntary registries [9].

#### High-risk and prioritized groups

Recruitment resources were focused on individuals who were most at risk from lead exposure during the FWC and who would greatly benefit from FR enrollment and referral to secondary prevention resources. High-risk subgroups included all children who were less than 18 years old during the FWC, especially children less than 6 years old, children less than 6 years old with blood lead level greater than 5mcg/dL, and those exposed prenatally. Occupants of Flint addresses with lead or galvanized service lines were also considered a high-risk lead exposure group.

### Participant Recruitment Sources

#### List-based recruitment

The FR recruitment plan included both a ‘self-referred’ sample (enrollees who respond to marketing and outreach) and a ‘list’ sample (enrollees who are recruited from population-based lists). Lists of individuals included names and contact information obtained through the CDC grant of public health authority or by individuals’ consent (Table 2). Through the public health authority designation, the FR requested and received permission from MDHHS to receive state program information (name, date of birth, address) of individuals with addresses corresponding to the City of Flint water system, for Medicaid, the Michigan Care Improvement Registry (MCIR), and the Childhood Lead Poisoning Prevention Program (CLPPP). The FR also requested and was approved by Hurley Medical Center, a public non-profit hospital in Flint, to receive contact information of patients with Flint city addresses. The FR requested to receive student contact information (student directory information) from Flint Community Schools and the GISD in compliance with public health legal or Family Educational Rights and Privacy Act (FERPA) exceptions. However, it was determined by both organizations that the Registry’s public health authority was insufficient to satisfy organization-specific policies governing release of this education directory data. Details on the public health and education laws that were considered are reported in a publicly available public health registry legal handbook [22]. Lists were also provided by local organizations not governed by the public health authority.

**Table 2.** Individual-level recruitment data sources for the Flint Registry.

| **Data Partner** | **Primary Client Base** | **Consent Requirements** |
| --- | --- | --- |
| MDHHS – Michigan Care Improvement Registry | Database of all individuals born in the state of Michigan or who received immunization care in Michigan | Data available through CDC Grant of Public Health Authority |
| MDHHS – Medicaid | State of Michigan Medicaid enrollees | Data available through CDC Grant of Public Health Authority |
| MDHHS – Childhood Lead Poisoning Prevention Program | Michigan children with blood lead test results | Data available through CDC Grant of Public Health Authority |
| Hurley Medical Center | Persons who utilized Hurley clinics, emergency departments, and in-patient care facilities | Data available through CDC Grant of Public Health Authority |
| CRIM Fitness Foundation Community Health Workers | Flint area school families and community members | Data available with client consent |
| Genesee Health System | Flint community members seeking mental health services | Data available with client consent |
| Greater Flint Health Coalition | Flint community members seeking health insurance and connection to a medical home | Data available with client consent |

Community outreach partners from organizations focused on assessing family needs, providing mental health services, and connecting community to medical services with clients in the Flint area consented individuals to share contact information with the FR for the purpose of recruitment (Table 2). As these individual-level data are received, they are deduplicated and used for participant recruitment lists (Table 3). All recruitment data are stored on HIPAA-compliant servers and access is restricted. A list of Flint city households with lead or galvanized service lines were sent specialized recruitment mailings (https://FlintPipeMap.org). Finally, comprehensive city-wide residential mailing lists are used.

**Table 3.** Household-level recruitment data sources for the Flint Registry.

| Data Source | Content |
| --- | --- |
| BlueConduit – FlintPipeMap.org | Database of Flint homes and service line composition |
| City of Flint Address Database | Commercially available list of city of Flint residences |

#### Community-outreach-based recruitment

Our community outreach recruitment strategy includes connecting FR staff or community partners directly with potentially eligible participants at local events. The FR engagement and enrollment workgroup includes community liaisons from programs that provide services to mitigate the effects of lead exposure. These liaisons were funded to integrate FR outreach scripts and materials into their established community activities. The FR Director of Community Engagement and Implementation and the Outreach Coordinator build community trust and use a diversified community-engaged outreach approach to support the FR with participation at both virtual and in-person events. FR outreach staff were present at recurring locations throughout the city and at FR outreach events geared towards high-priority audiences. In addition, the FR provided training for multi-sector FR Community Ambassadors. Ambassadors are recruited from community organizations and by using newsletters and social media. All ambassadors receive ongoing newsletters, which share upcoming FR events and opportunities for involvement, as well as community-based events, volunteer and donation needs, and upcoming ambassador training dates to be shared among the ambassadors’ various networks.

#### Marketing-based recruitment

Marketing-based recruitment is ongoing within the overall FR communication plan. The communication plan is designed to encourage ongoing awareness and enrollment in the FR (Figure S1), disseminate key findings to community, and maintain contact with existing enrollees. This marketing-based process is done in concert with and works alongside communications, outreach and engagement to extend and deepen the recruitment arm of the FR. Marketing efforts also support lead-specific education and training through distribution of a FR-supported Flint Lead Free report and other lead training events. Marketing recruitment efforts began broad and, as the FR matures, have transitioned to focused campaigns to further engage high-risk and difficult-to-reach populations for enrollment. The FR communications plan and messaging strategy was informed by the communications workgroup with experts in health communication, health literacy, advertising, media, and cultural competence and led by a Communications Director. Marketing materials have included commercials, billboards, bus signs, mailings, resident testimonials, and other promotional materials. Printed materials were provided in multiple languages, and video materials included both spoken English and adult sign language. Marketing and communications have been further enhanced through a social media presence and person-to-person outreach. The FR website is an easy-to-navigate, ADA- and WCAG-compliant, and mobile-friendly digital space to foster communication with registrants, provide registry progress updates, and summarize key findings. Regular newsletters are sent to enrollees to maintain updated contact information and to share results.

#### Recruitment protocol

Individuals recruited through list-based methods received invitation and reminder mailings presenting two methods of enrollment: 1) an online survey code used to complete eligibility, consent, and enrollment surveys, and 2) a phone number used to complete surveys administered by a telephone interviewer (Figure S2). The FR website provides a web-based form to request a personalized link (text or e-mail) to complete eligibility, consent, and the enrollment survey. Individuals learning about the project through marketing or outreach are directed to the website or provide contact information directly to outreach staff. Interview staff are available via a toll-free number to answer questions and complete eligibility screening, consent, or the enrollment survey over the phone. Staff also make outgoing calls to recruit and enroll participants and send texts and e-mails to encourage potentially eligible individuals to enroll. High-risk individuals receive additional focused mailings, text messaging, and phone calls. Focused recruitment with partner organizations was employed to locate and recruit high-risk individuals. For example, children with blood lead levels > 5 mcg/dL were part of the GFHC’s elevated blood lead level nurse case management program. Leveraging a pre-existing relationship with these families, GFHC staff connected with the children/families to further encourage FR enrollment.

### Survey Design Process

Our assessment workgroup developed the enrollment survey in collaboration with our community advisories with goals of creating an instrument sensitive to measuring health issues related to the water crisis and to collecting data to allow screening for referrals to services. The first step was an extensive review of existing large, epidemiological studies for relevant, national, validated survey items (e.g. National Health and Nutrition Examination Survey, Behavioral Risk Factor Surveillance System, Pregnancy Risk Assessment Monitoring System, National Survey of Children’s Health, World Trade Center Health Registry, and others) to allow for comparison of findings to other large studies (Table 4). Core data collected at enrollment includes demographics, physical and mental health, child development milestones, prior utilization of services (timing, frequency, and types), and environmental/lead-exposure risk screening. Screening tools for caregiver-reported child behavioral health (Behavior Assessment System for Children, 3rd Edition (BASC-3) and Behavior Rating Inventory of Executive Function, 2^nd^ Edition (BRIEF2)) were selected by the community workgroup, which included pediatricians, community mental health providers, local and county school district representatives, and child mental health researchers. Members of the CAB and Parent Partners piloted surveys, and content was adjusted based on community feedback.

**Table 4.**
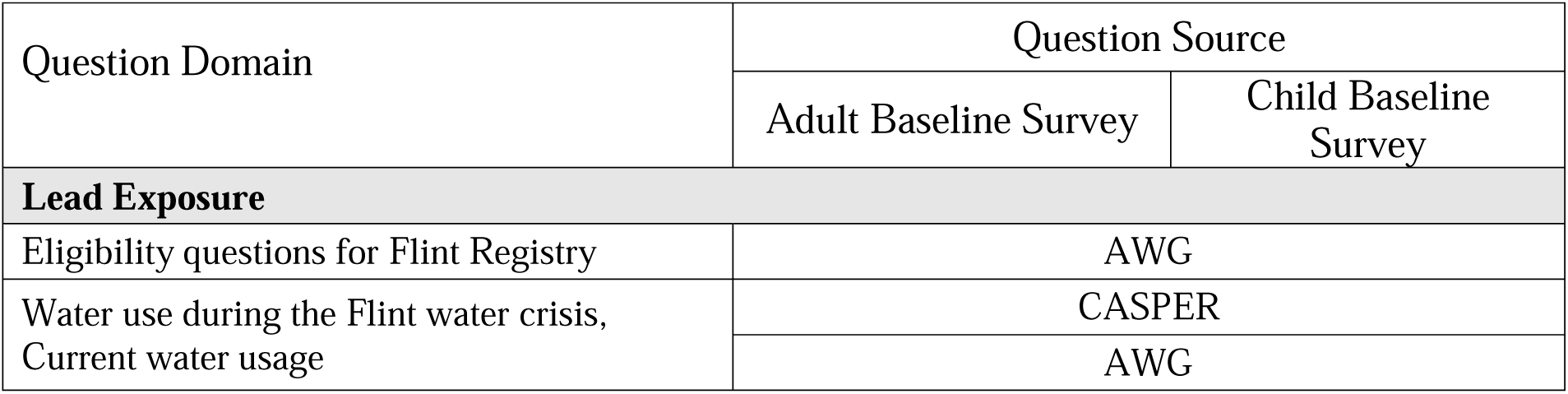
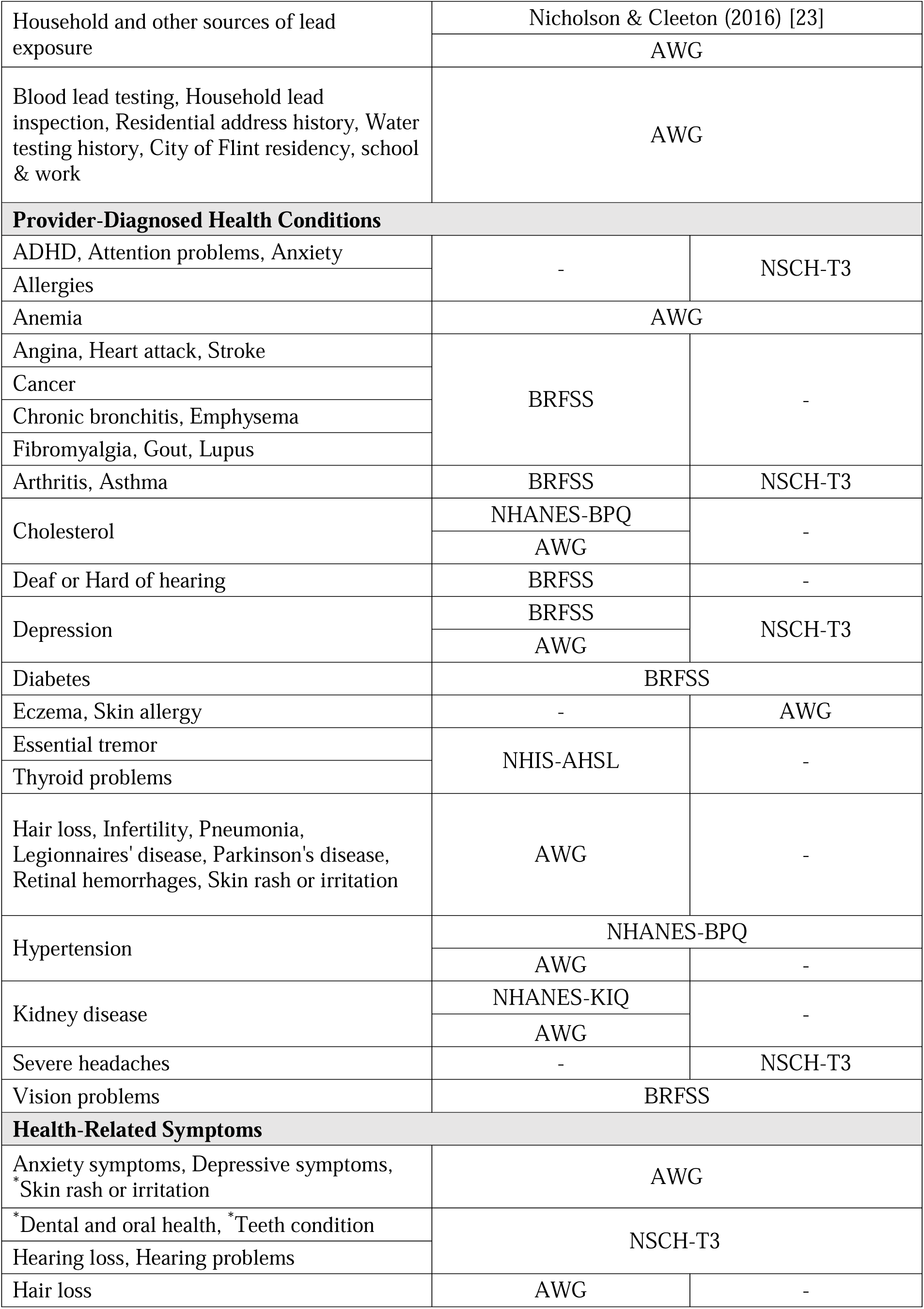
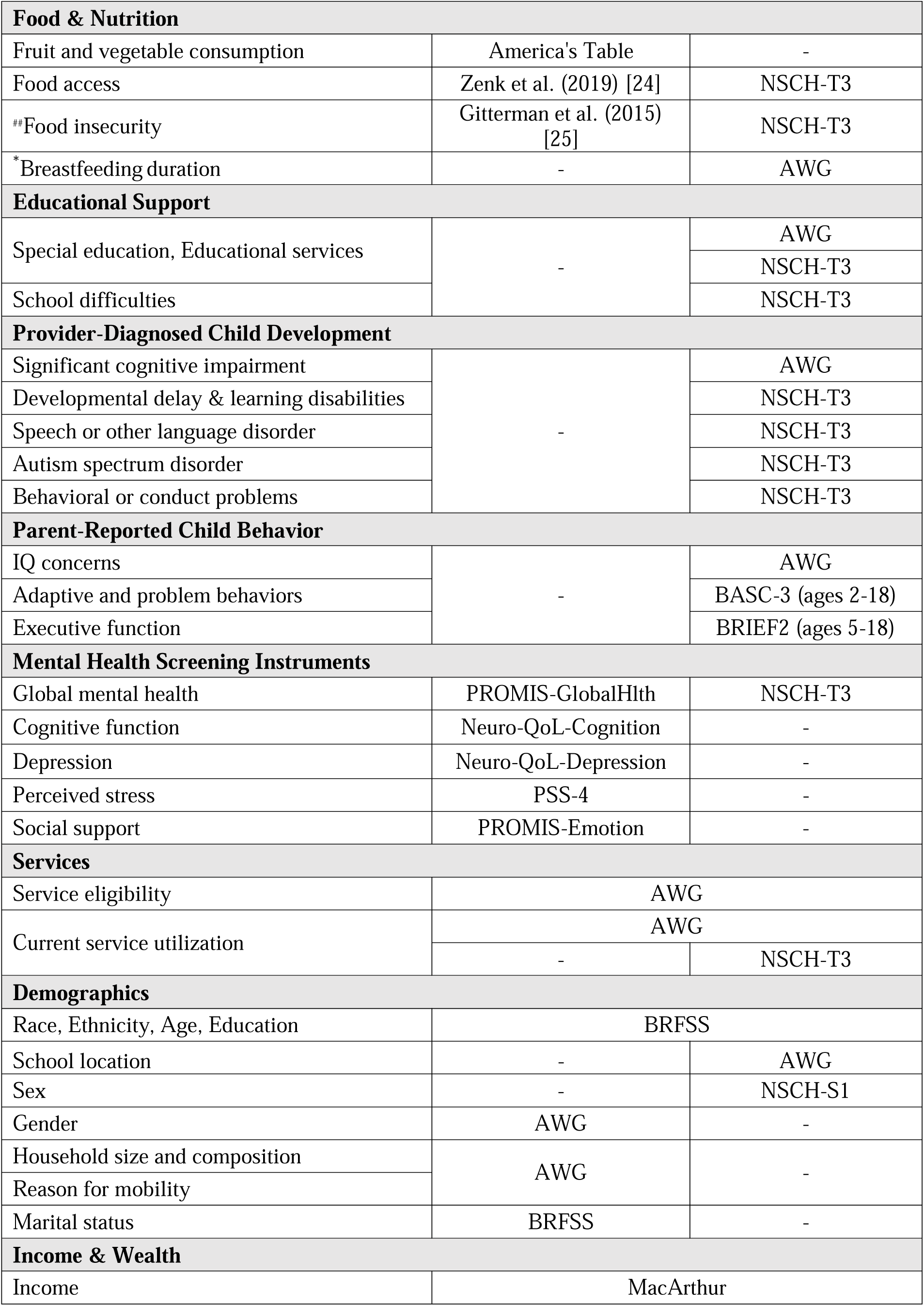
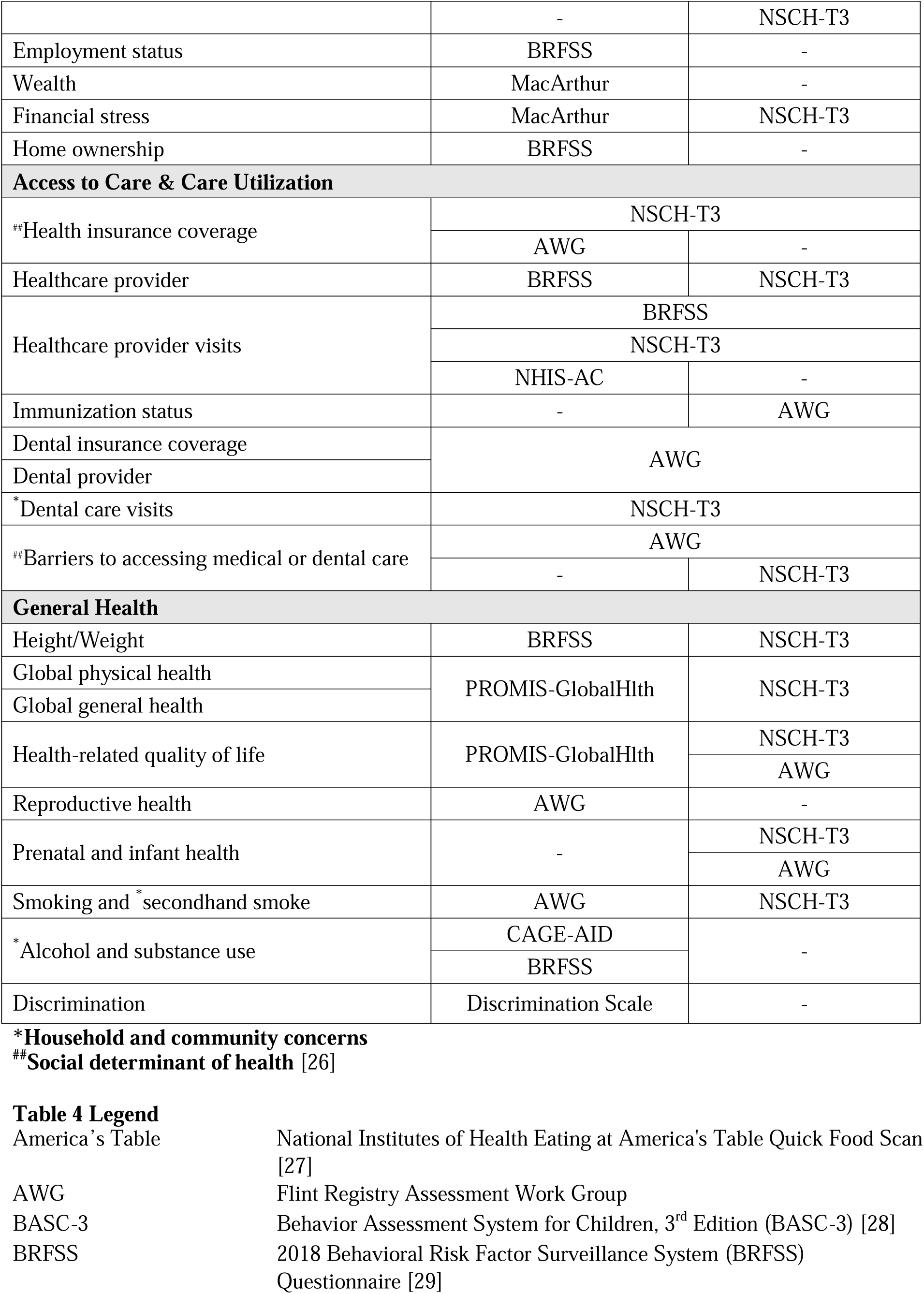
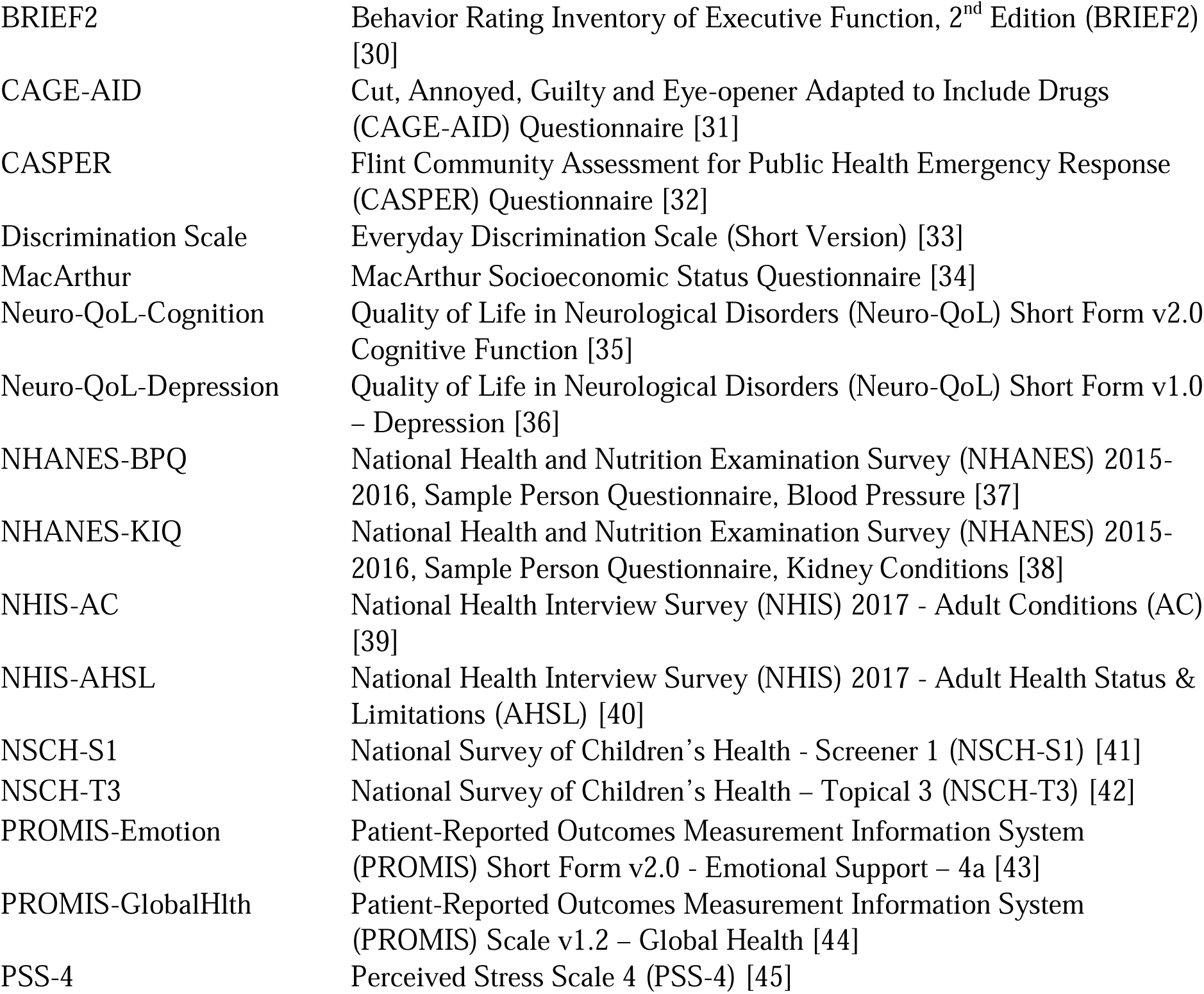
Flint Registry enrollment survey questions: domains and sources.

### Intervention Description

#### Referral network

The FR partnered with the GFHC to join and expand a multi-sector community referral network originally established under the GFHC’s Centers for Medicare & Medicaid Services–funded State Innovation Model Project. As part of this network, over 30 agency partners (Appendix A) are convened bi-monthly by GFHC to share best practices and emerging needs related to linking residents to resources and service providers that improve health outcomes. This network created a strong base of community partners focused on addressing social determinants of health and dedicated to tracking service referral outcomes across the community.

#### Referral process

Based on enrollment survey responses, registrants are screened and referred within the network to services to mitigate the effects of lead exposure. Referral screening is designed to be highly sensitive to limit the possibility of missing addressing participant needs. FR referrals are made to 14 agencies supporting 30 services in the categories of lead elimination, health, nutrition, and child development (Table 5). Three tiers of referrals are defined: 1) indirect referrals provide information to the FR participant about an available service, 2) direct referrals are sent on behalf of a participant to organizations via the Community Referral Platform (CRP-Software), and 3) supported referrals connect participants to a FR-supported MDHHS eligibility specialist to assist them in enrollment in MDHHS programs.

**Table 5.**
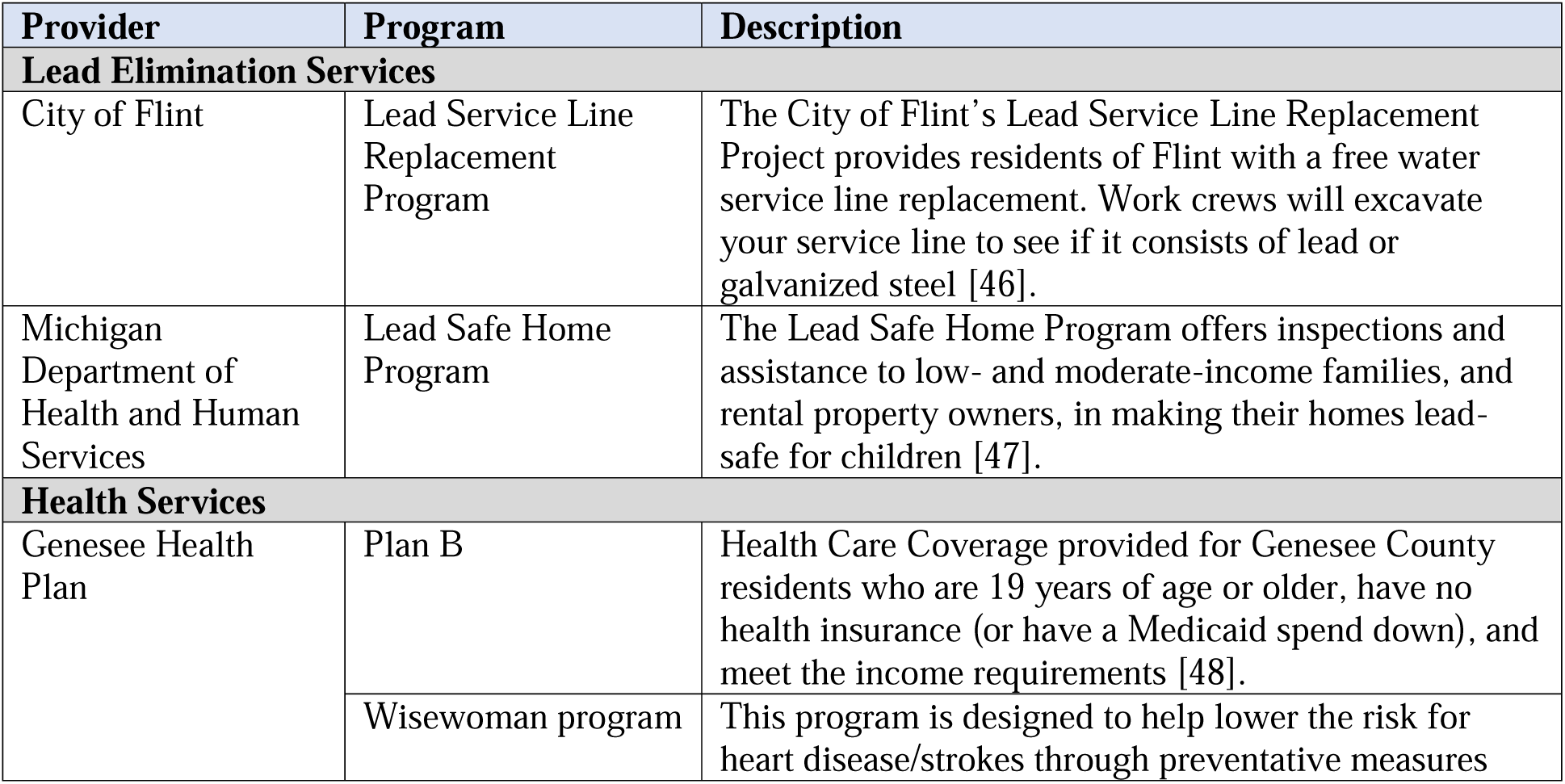
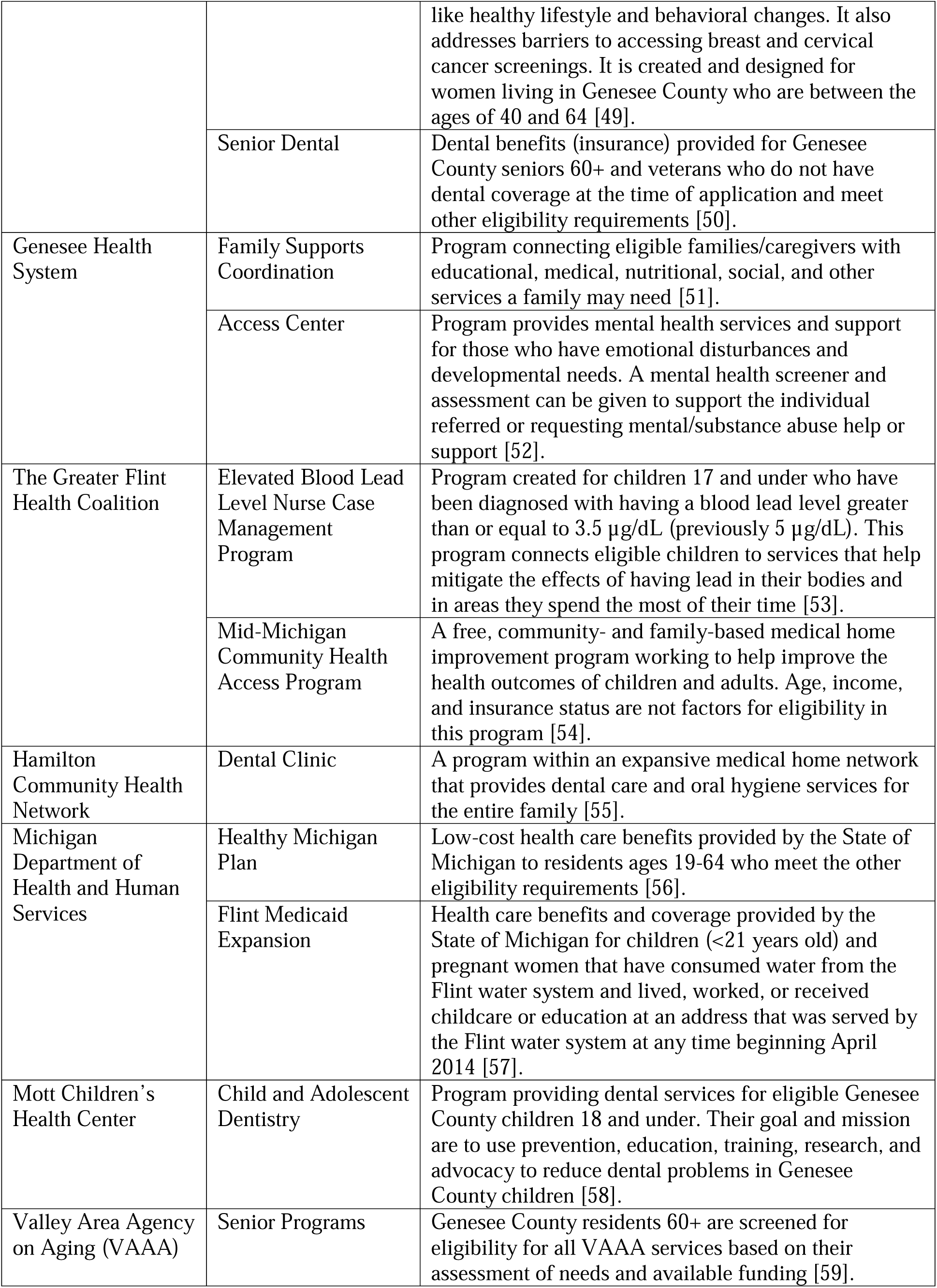
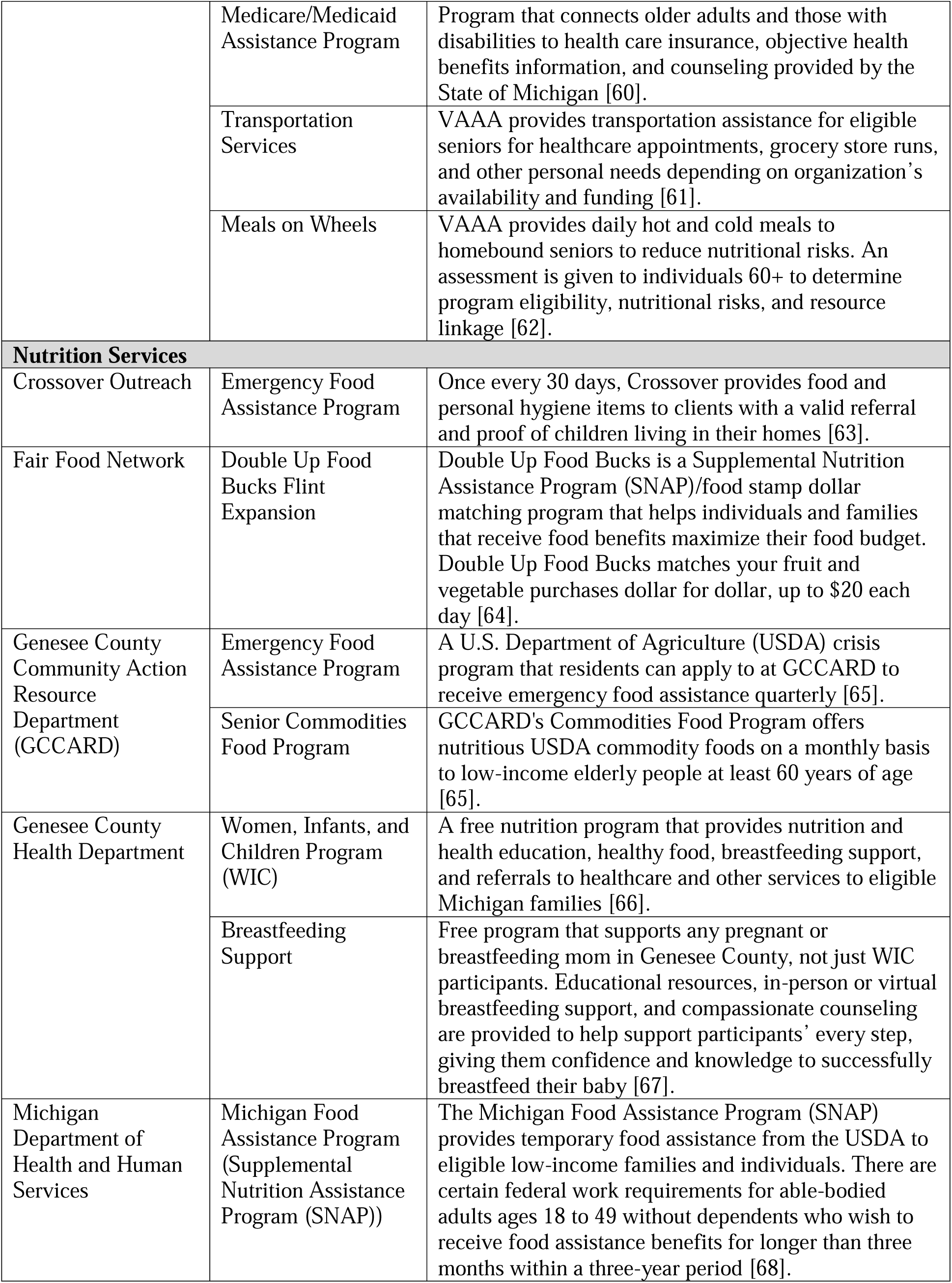
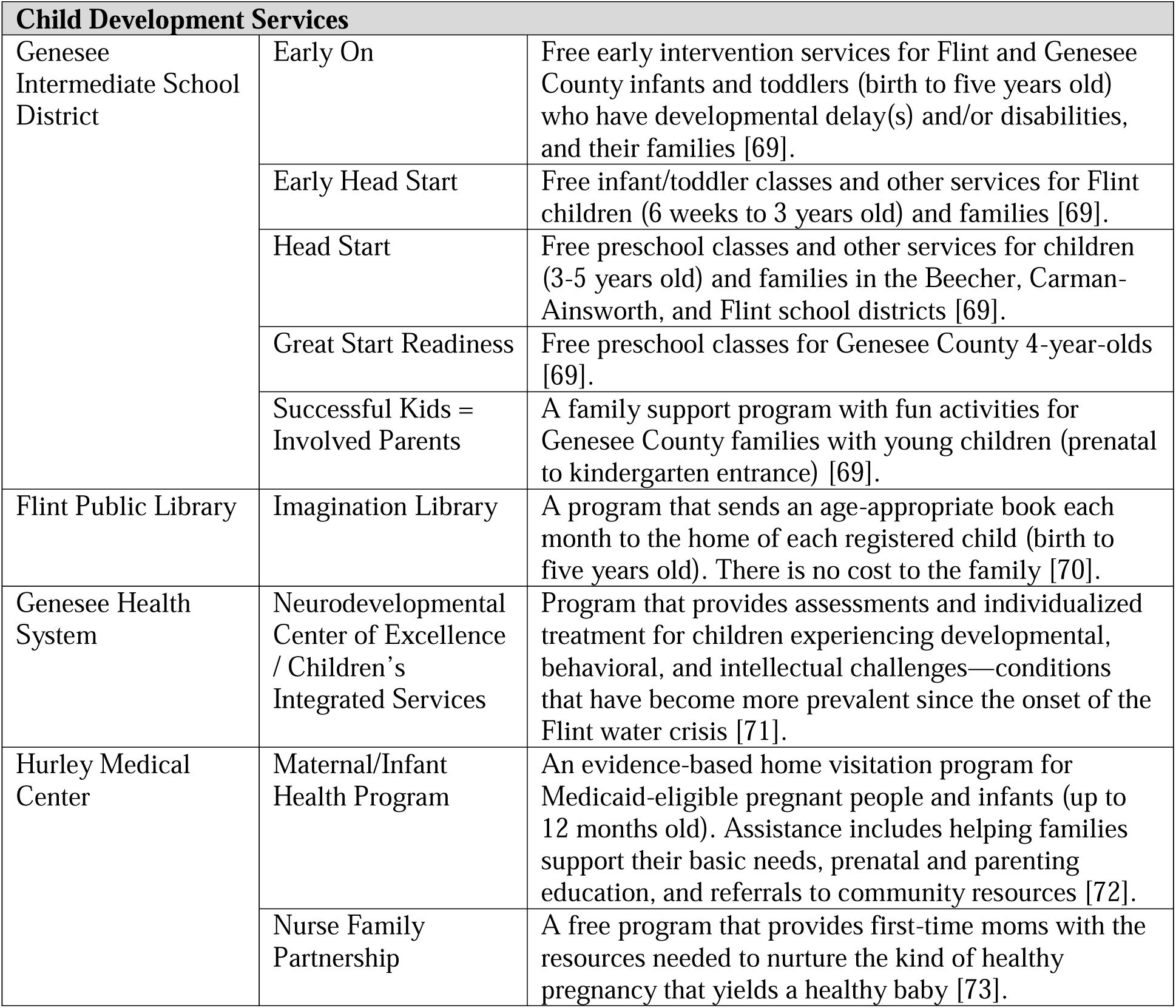
Service providers and programs, by Flint Registry referral domain.

### Data Collection Procedures

#### Staff and training

A well-trained, knowledgeable staff is important for the overall quality of the FR and for building trust with community partners and participants. To ensure that data are handled confidentially and securely, all FR personnel complete training in relevant modules of NIH Good Clinical Practice for Social and Behavior Research [74] and in FR-specific processes of data confidentiality and security. Personnel receive initial and ongoing education in environmental lead health and safety to ensure they are generally informed about the hazards of environmental lead exposure and the challenges facing those impacted by the FWC. Staff also receive training in trauma-informed responses. To help achieve high-quality data collection and minimize interviewer bias, interviewers receive initial training in Response Sensitive Standardized Interviewing (RSSI, University of Wisconsin Survey Center). Interviewers also receive ongoing retraining and certification along with quality checks and blinded telephone observations to ensure adherence to study protocols.

#### Data systems

Three software platforms are used to support key operational domains of the FR protocol: participant communications, survey data collection and storage, and participant referrals. An instance of the Research Electronic Data Capture system (REDCap) [75,76] maintained at Michigan State University (East Lansing, MI) is used to deploy participant surveys and store survey data. REDCap is a web-based application optimized for human subject research with robust data management tools and methods for secure data transfer. Eligibility, consent, and enrollment surveys are administered to participants via REDCap. An instance of Epic Systems maintained at Hurley Medical Center (Flint, MI) is used to de-duplicate participant records and drive completion of the FR recruitment protocol for each participant. Basic demographic information and a subset of key survey administrative variables are copied from REDCap to Epic via an HL7 interface [77]. Epic deduplication functionality is used to identify and exclude identical subject records that may arise from the open web enrollment survey format deployed via REDCap. Participant communications, sending of email, text, printed letter, and survey completion dates, are tracked using Epic reporting features.

Michigan Health Information Network (MiHIN) provided the technology to facilitate the referral process via its established CRP-Software. The CRP-Software was already in use in the Flint community, and leveraging its use to make FR referrals reduced the burden for service providers who were familiar with it. The CRP-Software is a closed-loop referral system and functions much like email, including participant-specific tracking functions, Accessed by both referral sender and recipient, to support efficient communication and follow-up for each referral made. The FR utilizes CRP-Software to track referrals made, received, and completed.

#### Eligibility screening, consent, and enrollment survey

Interested adults are asked to report if they lived, worked, or went to school in the city of Flint from April 25, 2014, to October 15, 2015. Adult parents or guardians complete eligibility, consent, and enrollment surveys on behalf of their minor children. Children born prior to August 1, 2016, are eligible if they lived, worked, went to school or daycare in the city of Flint from April 25, 2014, to October 15, 2015. The FR public health law team reviewed and managed legal and ethical consent issues and assisted in the development of a granular consent process that enables participants to preferentially consent to the following: to participate in the FR, to share eligibility and contact information with referral agencies, to be contacted for future research, and to share data from MDHHS back to the FR for outcomes evaluation. To reduce the barriers of technology and literacy to participants, the FR enrollment survey is offered in multiple modes of access: telephone interview, in-person interview, mailed paper-edition survey, and online survey (Table S1). Participants receive a bank check of $50 as a token of appreciation for their time and effort spent completing the enrollment survey.

### Exposure Measures

Exposure data are collected to determine eligibility and to further understand the extent of the crisis. Responses to eligibility questions determine whether individuals were exposed due to employment, residency, daycare, or school attendance. Additional details are collected about the length of time individuals were employed, went to school, or lived in the city from April 25, 2014, to October 15, 2015. Responses to survey questions describe the frequency of usage of unfiltered tap water (every day, less than every day, not at all, or unknown) during the FWC for six different categories of usage (drinking, washing dishes, brushing teeth, cooking, bathing/showering, and other). Residential address history within the city will also be used to further understand exposure related to service line composition. Data related to timing, frequency, type of exposure to the FWC, along with residential lead information will be combined to create a lead-in-water risk score to estimate the intensity of exposure.

### Surveillance and Mitigation Measures

Surveillance related to exposure is conducted by measuring the frequency (prevalence) and timing of health diagnoses, symptoms of health issues, nutrition access and insecurity problems, educational support needs, and child development diagnoses (Table 4). In addition, summary scores are calculated using mental health screeners and parent-reported child behavioral health questions in the BASC-3 and BRIEF2 (ages 5–18) or the BASC-3 only (ages 2–5) instruments (Table 4). Global health is measured for general health, physical health, mental health, and health-related quality of life. The total number of referrals made to each service provider and within each service domain are counted to describe health needs. The outcomes of FR referrals made for participants are tracked (completed by service provider, participant declined service, participant was not eligible for the service they were referred to, service provider could not contact participant) in each secondary prevention category to assess the status of the referral process.

### Covariates

Covariates are included in surveys to describe the characteristics of FR enrollees, as important predictors of overall health, or to use for risk stratification. Demographics include race, ethnicity, age, education level, gender or sex, household size, household composition (number of adults), and marital status. Economic measures include income, employment status, personal savings, hardship, and home ownership. Individual health characteristics include reproductive health, prenatal health, breastfeeding duration, fruit and vegetable consumption, smoking and secondhand smoke exposure, alcohol/substance use, and experiences with discrimination.

### Statistical Analysis Plan

The distributions of demographics in city of Flint enrollees will be compared to those from census data to describe the representativeness of the FR as compared to the city of Flint. Frequencies of reported health diagnoses, symptoms of health issues, nutrition access and insecurity problems, educational support needs, and child development diagnoses will be compared to city, county, state, and national measures as available to identify ongoing disparities. Child behavioral health scores from the BASC-3 and BRIEF2 will be compared to standardized scores to further describe child development disparities. Total referrals made in each domain (lead elimination, health, nutrition, child development) will be used to describe overall secondary prevention service needs (Table S2). Additional analysis will be conducted based on investigator-initiated hypotheses.

## DISCUSSION

The creation and implementation of a public health exposure registry in response to the FWC posed several challenges. First, the FWC was a prolonged crisis without a timely and direct measure of exposure in the participant and/or the participant’s environment. Second, solely conducting surveillance was not an option when considering the potential exposure sequalae. Third, the Flint community suffered from acute and chronic injustices with lost trust in institutions and government. Fourth, the denominator was massive, and the scope of referral services was broad.

The protocol developed for the FR was able to successfully solve these challenges. The protocol employed an inclusive and broad definition of exposure with tiered layers of risk populations. The FR protocol was designed to reach city-wide participation, with maximal flexibility, multiple modes of entry, and minimal barriers to enrollment.

The FR protocol is unique in that the FR conducts longitudinal surveillance and improves public health through a community-wide referral process. This strategy was determined to be the most appropriate to provide secondary prevention resources to mitigate the potential long-term manifestations of the water crisis. In implementing the referrals, the FR benefited from existing infrastructure such as a community referral network, a community referral software platform, and established community partnerships.

There were some challenges to this approach. By relying upon an existing system, the data collected by referral agencies were not consistent across all partners and not all partners were able to share individual-level data on referral dispositions. One ongoing concern related to the automated referrals is if participants will utilize services based on the online referral system. The feedback system of the CRP-Software has allowed us to monitor outcomes of referral to adjust referral criteria and outreach and communication strategies. Another challenge of the referral protocol is it is unknown which services will be referred to the most over time and therefore what capacity partner referral organizations need to support the FR. To address this concern, the FR needs to be prepared to bring on additional referral partners if capacity exceeds existing partner resources.

Another logistical challenge faced by the FR included utilizing an open survey link to allow enrollment in response to outreach and marketing. While this allowed enrollees the ability to sign up and quickly complete surveys, it required time-intensive data management activities to remove duplicate enrollees and to verify participant identities.

Most important of all, recognizing the community-wide trauma and betrayal, the FR protocol was developed in partnership with community and significant resources were invested in community engagement to build trust and include diverse community voices.

The FR protocol is applicable to other public health crises in several ways: The eligibility criteria were developed with no direct measure of exposure which is often the case in environmental health exposures, surveillance and mitigation efforts needed to be broad and conducted on a city-wide scale, the protocol was designed with flexibility to reduce barriers and maximize participation, and community engagement and partnership were paramount to its success.

The future work of the FR will include ongoing enrollment and referrals along with long-term surveillance and mitigation efforts. Linkages to other health and environmental data will allow for further assessment of the impact of the FWC. Data collected as part of future surveys will allow for evaluation of the referral process. Dissemination efforts will include academic, community, and government audiences to share lessons learned, policy implications, and ongoing community needs.

## Data Availability

Requested data may be provided after IRB approval and appropriate data use agreements have been obtained.

## LIST OF ABBREVIATIONS

FWC: Flint water crisis
CDC: Centers for Disease Control and Prevention
FR: Flint Registry
MSU: Michigan State University
GFHC: Greater Flint Health Coalition
MDHHS: the Michigan Department of Health and Human Services
CAB: Community Advisory Board
CERB: Community Ethics Review Board
ACS: American Community Survey
GISD: Genesee Intermediate School District
MCIR: Michigan Care Improvement Registry
CLPPP: Childhood Lead Poisoning Prevention Program
BASC-3: Behavior Assessment System for Children, 3^rd^ Edition
BRIEF2: Behavior Rating Inventory of Executive Function, 2^nd^ Edition
ADHD: Attention-deficit/hyperactivity disorder
CRP: Community Referral Platform
VAAA: Valley Area Agency on Aging
SNAP: Supplemental Nutrition Assistance Program
GCCARD: Genesee County Community Resource Department
USDA: U.S. Department of Agriculture
WIC: Women, Infants, and Children
REDCap: Research Electronic Data Capture

## DECLARATIONS

## Ethics Approval and Consent to Participate

The Michigan State University Institutional Review Board reviewed the FR protocol and determined it did not meet the definition of “research” as defined by the U.S. Department of Health and Human Services regulations for the protection of human research subjects. In addition, the FR received an endorsement from the local volunteer-led Community Based Organization Partners Community Ethics Review Board (CERB) after a complete review which determined the Flint Registry project to be “ethically sound and consistent with the vision and mission of the CERB.” Adults provide consent to participate in the FR, and adult parents or guardians provide consent on behalf of their children.

## Consent for Publication

Not applicable

## Authors’ Contributions

All authors contributed to the conception and design of the work, drafted significant portions of the manuscript, contributed to the interpretation of data, and read and approved the final manuscript. NJ and MC made substantial contributions to the acquisition of data. All authors are personally accountable for their own contributions and ensure that questions related to the accuracy or integrity of any part of the work, even ones in which the author was not personally involved, are appropriately investigated, resolved, and the resolution documented in the literature.

## Acknowledgements

The authors thank Chris Hippensteel for their denominator work, Imari Smith for documenting service providers, Sarah Jenuwine for summarizing supplemental data, as well as Katherine Negele and Katlin Harwood-Schelb for their assistance with manuscript preparation.

