## Appendix A for "Study Protocol for Developing a Public Health Registry: The Flint Registry Experience"

**Community Referral Network Partner Agencies**

1. Catholic Charities of Shiawassee and Genesee Counties
2. Child Care Network
3. City of Flint
4. Crossover Outreach
5. Easterseals of Michigan
6. Food Bank of Eastern Michigan
7. Mid-Michigan Community Health Access Program
8. Genesee Health System
9. Genesee Intermediate School District
10. Greater Holy Temple
11. Michigan State University
12. My Brother’s Keeper
13. New Paths, Inc.
14. Sacred Heart Rehabilitation Center
15. Salvation Army
16. United Way of Genesee County
17. Valley Area Agency on Aging
18. Carriage Town Ministries
19. Family Service Agency
20. Flint Public Library
21. Genesee County Community Action Resource Department
22. Genesee County Health Department
23. Hamilton Community Health Network
24. Latinx Technology and Community Center
25. Legal Services of Eastern Michigan
26. Mass Transportation Authority
27. Michigan Health Information Network Shared Services
28. Shelter of Flint
29. The Disability Network
30. Traverse Place
31. YWCA of Greater Flint
