## Supplemental Data for "Study Protocol for Developing a Public Health Registry: The Flint Registry Experience"

**Figure S1. Percentage of enrollees who reported hearing about the Flint Registry by specific marketing and outreach methods (multiple responses allowed) through July 2022**

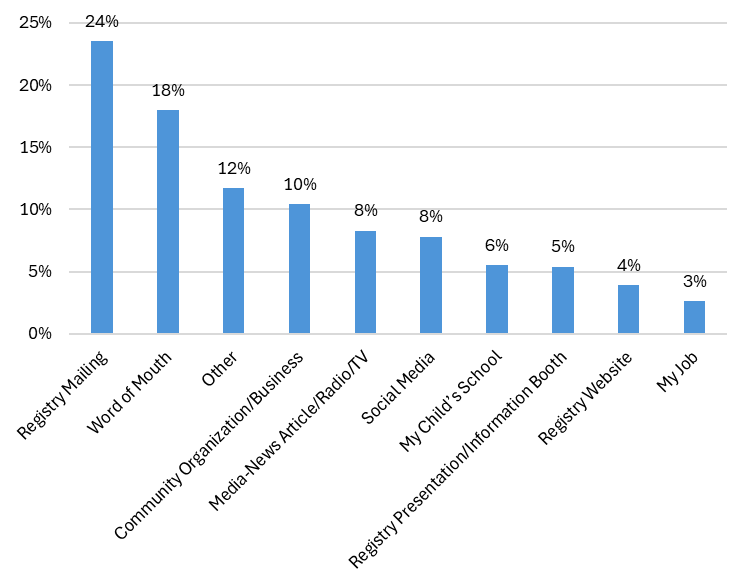

N = 20,379; Multiple Responses Allowed

**Figure S2. Flint Registry enrollment through July 2022**

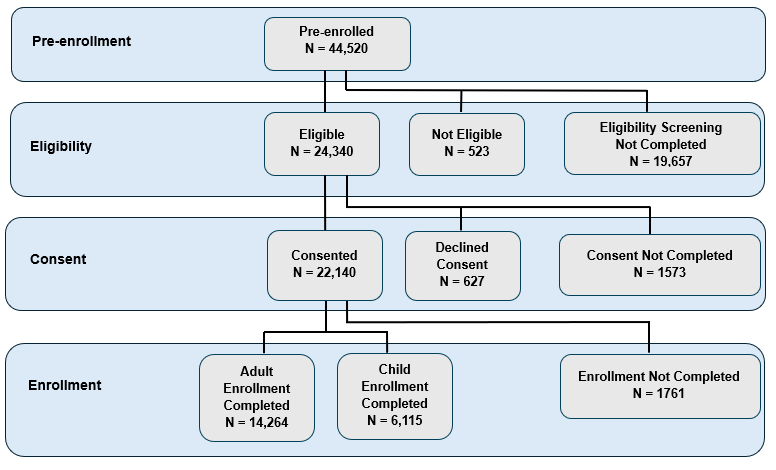

Pre-enrolled includes a database of potential participants that either indicated they were interested in response to outreach or marketing, or were identified as a high-risk or priority group and had contact information in a public health database (children who were less than 18 years old during the Flint water crisis, children less than 6 years old with blood lead levels greater than 5 mcg/dL, and those exposed prenatally). City-wide mailings may have engaged other potential participants that were not pre-enrolled.

Eligibility screening, consent and enrollment are three separate surveys. “Eligibility Screening Not Completed” indicates eligibility screening was not completed due to non-response during the follow-up period or the individual declined to complete the screening. “Consent Not Completed” and “Enrollment Not Completed” indicate these were not completed due to non-response during the follow-up period.

91.7% of enrollees were Flint residents (N=18,690) and represented 18.6% of the estimated population of exposed Flint residents (N=100,569).

**Table S1. Percentage of Flint Registry enrollees by survey administration mode through July 2022 (N=20,379)**

| **Mode of Survey Enrollment** | **Adult Survey (N=14,264)**  **%** | **Child Survey (N=6,115)**  **%** |
| --- | --- | --- |
| Online Survey | 69.6 | 80.7 |
| Phone Survey | 22.9 | 14.1 |
| Mailed Paper Survey | 5.2 | 4.1 |
| In-person Survey | 1.3 | 0.2 |
| Multiple Survey Modes | 1.0 | 1.0 |

**Table S2. Total and percentage of Flint Registry referrals sent by referral domain** **through July 2022**

| **Referral Domain** | **Total Referrals Sent** | **Percentage**  **of**  **Total** |
| --- | --- | --- |
| Lead Elimination Services | 8895 | 31.8 |
| Health Services | 7799 | 27.9 |
| Nutrition Services | 6801 | 24.3 |
| Child Development Services | 4449 | 15.9 |
| Total | 27,994 |  |
